# A STUDY TO ANALYSE THE DEMOGRAPHICS AND INJURY PATTERN OF DOG BITE CASES IN EMERGENCY DEPARTMENT OF A TERTIARY CARE HOSPITAL IN CHENNAI

**DOI:** 10.64898/2026.05.20.26353645

**Authors:** Dhanalakshmi Vinoth, A. Ashok Kumar, V. Evangelin Jenifer, S. Raj, C. Keerthana, K. Arthi

## Abstract

**Background:** Dog bite injuries are a major yet largely preventable public health concern worldwide. They contribute significantly to morbidity, healthcare burden, and economic costs, particularly in emergency department. The present study aims to analyse the demographics and injury pattern of dog bite cases presenting to the emergency department of a tertiary care hospital in Chennai.

**Methods:** We conducted a cross-sectional study with dog bite injured participants attending the Causality from November 2025 to April 2026 data was collected using a structured tool including details on demographics (Age, Gender, Education) injury related characteristics, history of pure bite site of dog bite injury type, WHO bitten criteria and information to management etc. We used here non probability statistical analysis and age specific dog bite cases and independent variables were analysed using SPSS (2.0 version).

**Result:** A total of Two hundred sixteen dog bite cases were analysed in the study by period of 6 months The majority of participants were 172 (79.6%) were above 18 years and 44 (20.4) were below 18 year, 130 (60%) from rural areas and 86 (39.8%) from urban areas, 136 (63.0%) of Victims presented within a day of the bite, 61(28.2) next day and 19 (8.8%) in after one week 66 (30.6) were bitten by own dog and 150(69.4%)were bitten by neighbour / friended dog. 124(57.4) were bitten by stray dog 92(42.6) bitten by pet dog. 117(54.2) were vaccinated dog and 99(45.8%) were not vaccinated. 110(50.9) victims were injured by laceration. 26(12.0%) were injured by puncture wound.46(21.3) were injured by abrasion 10(4.6) were injured by avulsion 15(6.9%) were injured by crush injury. 156(72.2%) were had minor wound.45(20.8%) victims had moderate wound and 15(6.9%) victims had severe wound. 112(51.9%) victims were taken antibiotics.104(48%) were not taken antibiotics. 185(85.6%) victims received tetanus toxoid, 31(14.4%) were not received tetanus toxoid.

**CONCLUSION:** There is a high burden of dog bite injuries from stray dogs in India. Despite early hospital presentation in many cases gaps in first aid practices and rabies post exposure prophylaxis were evident and highlighting inadequate awareness.

## 1. INTRODUCTION

Dog bite injuries are a major yet largely preventable public health concern worldwide. They contribute significantly to morbidity, healthcare burden, and economic costs, particularly in emergency department settings(Rabies, n.d.). Globally, dog bites account for tens of millions of injuries each year, although precise estimates remain unavailable due to underreporting and lack of surveillance systems in many regions(Animal Bites, n.d.-a). In high-income countries such as the United States, approximately 4.5 million dog bites occur annually, with hundreds of thousands requiring medical attention(Dixon et al., 2012). In contrast, data from low- and middle-income countries (LMICs) are fragmented; however, available evidence suggests that dogs are responsible for 76–94% of all animal bite injuries(Maniscalco et al., 2026)

The burden of dog bites is closely linked to rabies, a fatal zoonotic disease caused by a neurotropic virus of the genus Lyssavirus(Rabies, n.d.). Rabies remains a significant global health issue, causing an estimated 59,000 deaths annually, with over 95% of fatalities occurring in Asia and Africa, India alone accounts for a substantial proportion of these deaths, with approximately 18,000–20,000 fatalities reported each year(Rabies - India, n.d.). Despite the availability of effective vaccines and post-exposure prophylaxis (PEP), rabies continues to have a near 100% fatality rate once clinical symptoms appear(Rabies, n.d.).The higher mortality rates in LMICs are attributed to factors such as delayed treatment, poor awareness, limited access to healthcare, and the high prevalence of stray dogs(Chisare,D, 2023) Dog bites also impose a considerable strain on healthcare systems. In the United States, they account for approximately 337,000 emergency visits annually, with associated healthcare costs reaching up to $2 billion per year(Ortiz & Lezcano, 2023). Children are the most vulnerable group, often sustaining bites to the head and neck, while adults typically present with injuries to the extremities and hands(Animal Bites - StatPearls - NCBI Bookshelf, n.d.). Dog bites can lead to severe consequences, including serious physical injuries, frequent hospitalizations, potential infections, reconstructive procedures due to the potential for permanent disfigurement and psychological and post-traumatic stress(Khan et al., 2023). Appropriate wound management—including prompt cleaning, irrigation, and removal of devitalized tissue is essential to prevent complication(Manna et al., 2026). However, in many rural and underserved communities, traditional and unscientific practices such as the application of oils, herbs, or irritants to wounds persist, increasing the risk of infection and rabies transmission(Beasley et al., 2022).

Previous studies have highlighted the epidemiology and clinical patterns of dog bite injuries across different regions(Khan et al., 2023). Research from high-income countries has provided relatively reliable data on incidence, hospitalization rates, and mortality trends(English et al., 2018). For instance, studies in the United States and New Zealand have documented both fatal and non-fatal dog bite injuries, demonstrating an increasing trend in hospitalizations over time(Duncan-Sutherland et al., 2022). In LMICs, studies have primarily focused on rabies burden, awareness levels, and healthcare-seeking behaviour(Acharya et al., 2026). Indian studies emphasize the endemic nature of rabies, the predominance of dog mediated transmission (up to 97%), and the significant role of sociocultural practices in influencing treatment outcomes(Rabies - India, n.d.). However, many of these studies are community-based or focus on awareness rather than detailed clinical and demographic analysis in emergency settings(Garg et al., 2013).

Despite the substantial burden of dog bite injuries, there is a lack of comprehensive, hospitalbased studies analysing the demographic profile, injury patterns, and management practices of dog bite cases, particularly in urban tertiary care settings like Chennai(Singaravel et al., 2026). Additionally, limited data exist on how factors such as age, gender, injury site, and timing of presentation influence clinical outcomes. This gap hinders the development of targeted interventions and effective prevention strategies(Yattoo et al., 2009)

The present study aims to analyse the demographics and injury patterns of dog bite cases presenting to the emergency department of a tertiary care hospital in Chennai. By examining patient characteristics, injury distribution, and management practices, the study seeks to provide a clearer understanding of the burden and clinical profile of dog bite injuries in this setting. This study is expected to generate valuable data on the epidemiology and clinical characteristics of dog bite cases in an urban tertiary care setting. The findings will help identify high-risk groups, common injury patterns, and gaps in management practices. Ultimately, the study will contribute to improving emergency care protocols, enhancing public awareness, and guiding policymakers in designing effective dog bite prevention and rabies control strategies

## 2. MATERIALS AND METHODS

### STUDY DESIGN, POPULATION AND SETTING

We conducted a cross-sectional study with Dog bite injured patients presenting to ACS medical college and hospital, a tertiary care hospital in Chennai, India. Eligibility criteria included: dog bite victims of all ages visiting the dog bite unit of a significant public tertiary care hospital from November 2025 to April 2026

### SAMPLE SIZE CALCULATION

We used a non-probability, purposive sampling method to draw the sample. Data were collected and analysed from 216 patients with dog bite injuries. The sample size was calculated using the absolute precision formula, n = Z(α/2)^2^ P(1−P)/d^2^, considering a 95% confidence interval and 80% power of the study. Where Z(α/2)^2^ = 3.84, P = 0.765 (frequency and characteristics of dog bite injuries reported as 76.5%), 1−P = 0.235, and d^2^ = 0.0025 (precision 5.0%), the sample size calculated was 276. We added 10% to the calculated sample size to account for a non-response rate, giving a final sample size of 304. However, the sample size achieved was 216.

### STUDY PROCEDURE AND DATA COLLECTION

The study was conducted following a systematic and well-organized procedure to ensure the accuracy and consistency of the data collected. Prior approval was obtained from the concerned authorities, and the purpose of the study was clearly explained to all participants. Eligible participants were selected based on predefined inclusion criteria, and informed consent was obtained prior to their involvement, ensuring they were aware of their rights, including confidentiality and voluntary participation. Data collection was carried out by trained research assistants who approached dog bite patients receiving initial treatment and invited them to participate in the study. Information was gathered using a structured proforma, which included details on demographic characteristics (such as age, gender, and education), injury-related factors including date and time of injury, location at the time of the incident, history of previous bites, site and type of injury, whether the individual was alone or accompanied, circumstances and activity during the bite, and ownership status of the animal), as well as management-related aspects (such as first aid measures, hospital treatment, and type of vaccination received). Each participant was interviewed individually in a comfortable setting, and questions were explained in simple language to ensure clarity and accurate responses. Observations and available records were also used when necessary to supplement the data.

After data collection, all completed proforma were thoroughly reviewed for completeness and accuracy. The data were then organized, coded, and prepared for analysis, while maintaining strict confidentiality and adherence to ethical standards throughout the study. The exclusion criteria for the study included patients presenting with bites from animals other than dogs, such as cats, monkeys, snakes, rats, or humans; cases where injuries were not directly caused by dog bites (for example, scratches or injuries resulting from falls while being chased but not bitten); patients who received initial treatment elsewhere and presented only for follow-up without a documented record of the dog bite in the emergency department; cases with incomplete or missing medical records lacking essential study variables; and patients who refused to provide consent for participation.

### STATISTICAL ANALYSIS

Data will be entered into a secure spreadsheet (Microsoft Excel) and then imported into a statistical software package (e.g., SPSS version 2.0) for analysis. Descriptive Statistics: Categorical variables (gender, wound location, management type) will be presented as frequencies and percentages. Continuous variables (age) will be summarized using means and standard deviations or medians and interquartile ranges, depending on normality.Chi-square test (or Fisher’s exact test) will be used to assess associations between categorical variables (e.g., wound severity classification and bite location (head/neck vs. limbs).

## 3. RESULTS

**(TABLE -1& 2)** A total of 216 dog bite cases were analysed in this study by a period of 6 months. The majority of participants were aged above 18 years (79.6%, n = 172), while 20.4% (n = 44) were below 18 years. Males constituted 56.5% (n = 122) of cases, and females accounted for 43.5% (n = 94). Regarding educational status, 32% (n = 64) were educated and 76% (n = 152) were uneducated. Most participants were from rural areas (60%, n = 130), with 39.8% (n = 86) residing in urban settings. In terms of healthcare-seeking behaviour, 63.0% (n = 136) of victims presented within a day of the bite, 28.2% (n = 61) the next day, and 8.8% (n = 19) after one week. At the time of the incident, 46.8% (n = 101) were walking, 32.4% (n = 70) were playing or running roadside, and 20.8% (n = 45) were engaged in other activities. A history of provoking or playing with a dog was reported by 34.7% (n = 75), while 65.3% (n = 141) denied such actions; additionally, 17.6% (n = 38) reported intervening in a dog-related incident prior to the bite. Most bites were attributed to friends’ dogs (69.4%, n = 150), followed by neighbours’ dogs (30.6%, n = 66). The majority of incidents occurred on the street (50.9%, n = 110), followed by home (31.0%, n = 67) and workplace (10.2%, n = 22). Stray dogs were responsible for 57.4% (n = 124) of bites, while pet dogs accounted for 42.6% (n = 92). Attacks without provocation were reported in 43.5% (n = 94) of cases. Most dogs were described as friendly (64.4%, n = 139), while 35.6% (n = 77) were considered aggressive. Regarding vaccination status, 54.2% (n = 117) of dogs were vaccinated, and 45.8% (n = 99) were not; abnormal behaviour in the preceding 10 days was noted in 14.8% (n = 32). A single bite was reported in 87.0% (n = 188) of cases, while 13.0% (n = 28) experienced multiple bites; 17.6% (n = 38) of dogs had a previous history of biting. Following the incident, 75.9% (n = 164) of dogs remained alive, 9% (n = 2) died, and the status was unknown in 23.1% (n = 50). The lower extremities were the most commonly affected site (56.5%, n = 122), followed by upper extremities (38.0%, n = 82), with fewer injuries to the head and neck (2.3%, n = 5) and torso (3.2%, n = 7). Lacerations were the most common type of injury (50.9%, n = 110), followed by abrasions (21.3%, n = 46), puncture wounds (12.0%, n = 26), crush injuries (6.9%, n = 15), avulsions (4.6%, n = 10), and cellulitis (4.2%, n = 9). Based on severity, 72.2% (n = 156) of wounds were minor, 20.8% (n = 45) were moderate, and 6.9% (n = 15) were severe. In terms of management, 40.3% (n = 87) underwent primary suturing, 13.9% (n = 30) had delayed closure, 19.4% (n = 42) were left to heal by secondary intention, and 26.4% (n = 57) required no such intervention. Antibiotics were prescribed in 51.9% (n = 112) of cases. First aid measures included washing with soap and water (48.6%, n = 105), cleaning with antiseptic (31.0%, n = 67), and application of gel or ointment (20.4%, n = 44). Tetanus toxoid was administered in 85.6% (n = 185) of cases, while rabies post-exposure prophylaxis was given to 55.1% (n = 119). Hospital admission was required in 16.7% (n = 36) of cases, while 83.3% (n = 180) were managed on an outpatient basis, and 25.9% (n = 56) were referred to specialty clinics.

**TABLE -1.** SOCIO-DEMOGRAHIC.

| VARIABLES |  | FREQUENCY<br>(N=216) | PERCENTAGE<br>% |
| --- | --- | --- | --- |
| AGE | <18 YEARS | 44 | 20.4 |
|  | >18 YEARS | 172 | 79.6 |
| GENDER | MALE | 122 | 56.5 |
|  | FEMALE | 94 | 43.5 |
| EDUCATED |  | 64 | 32 |
| UNEDUCATED |  | 152 | 76 |
| AREA OF<br>RESIDENCE | RURAL | 130 | 60 |
|  | URBAN | 86 | 39.8 |

**TABLE -2.**
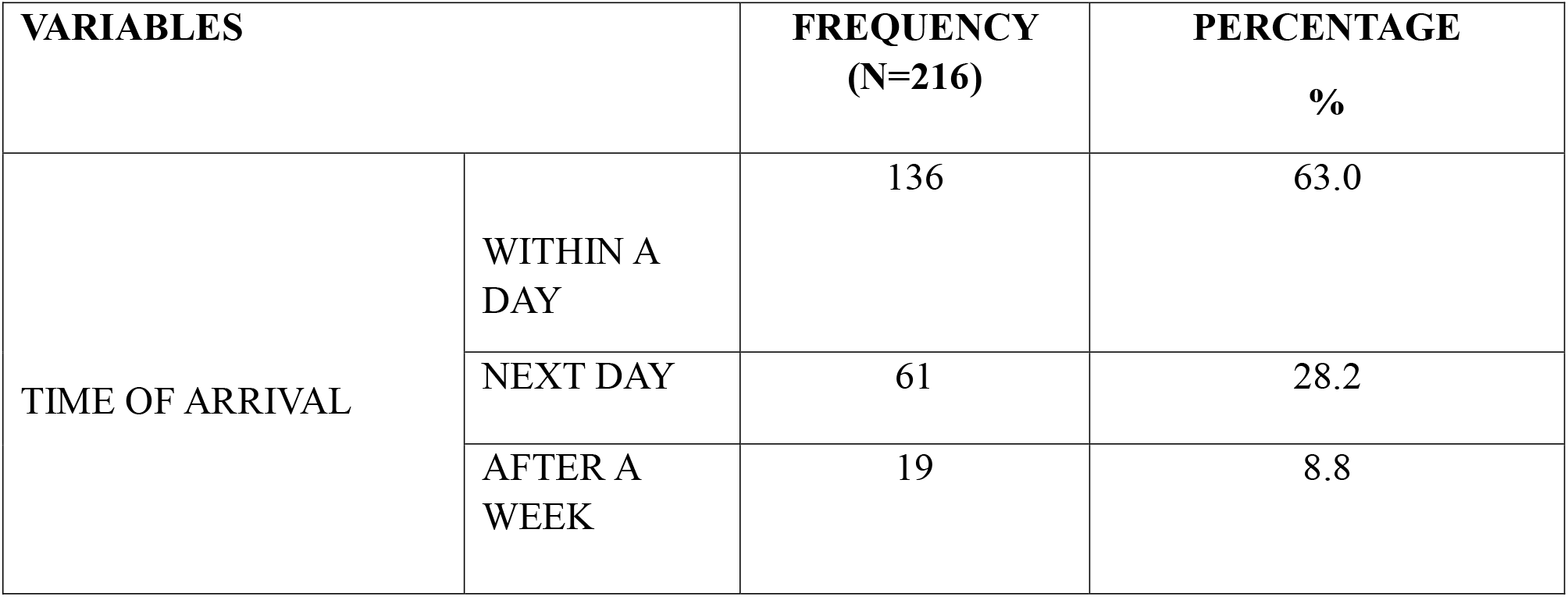
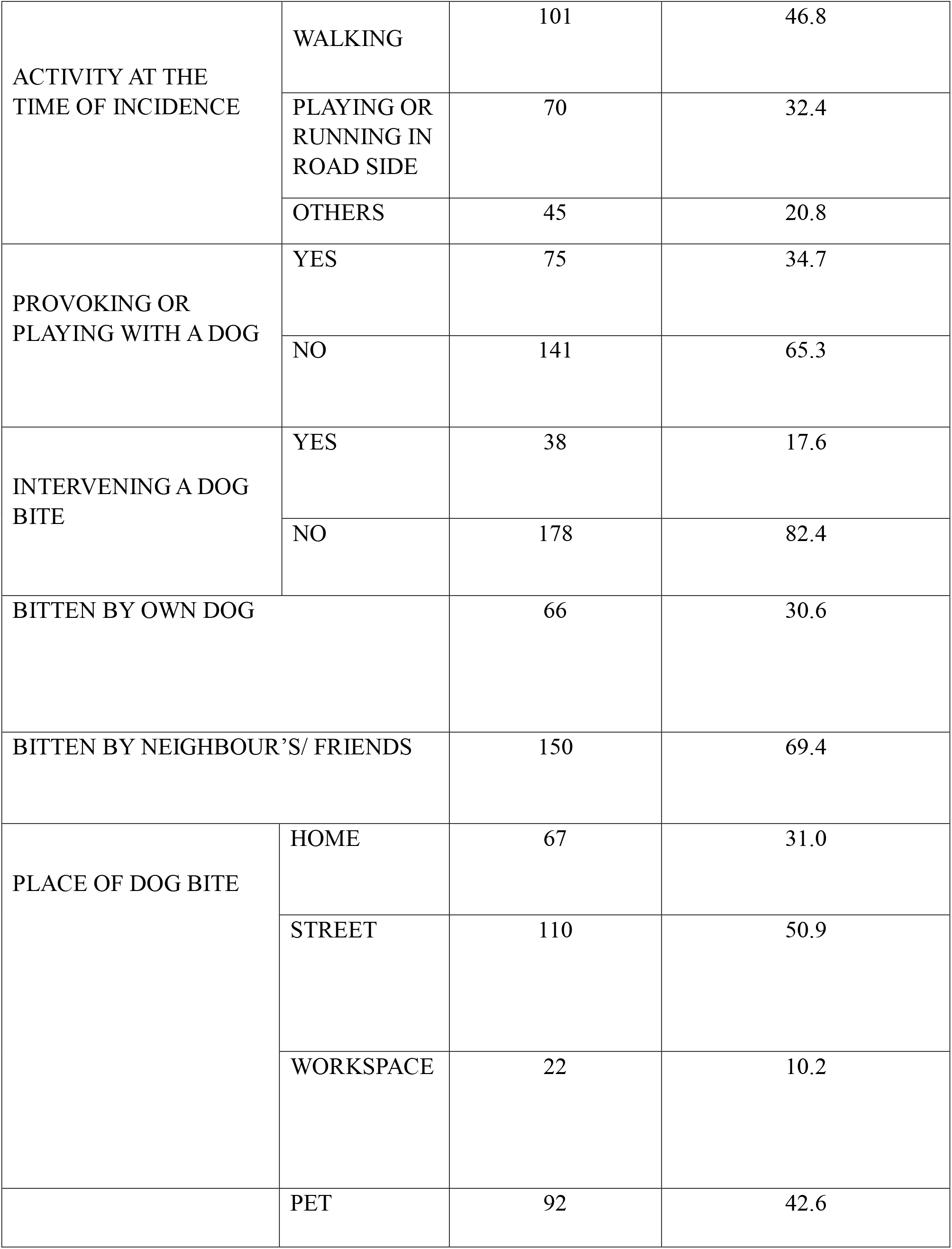
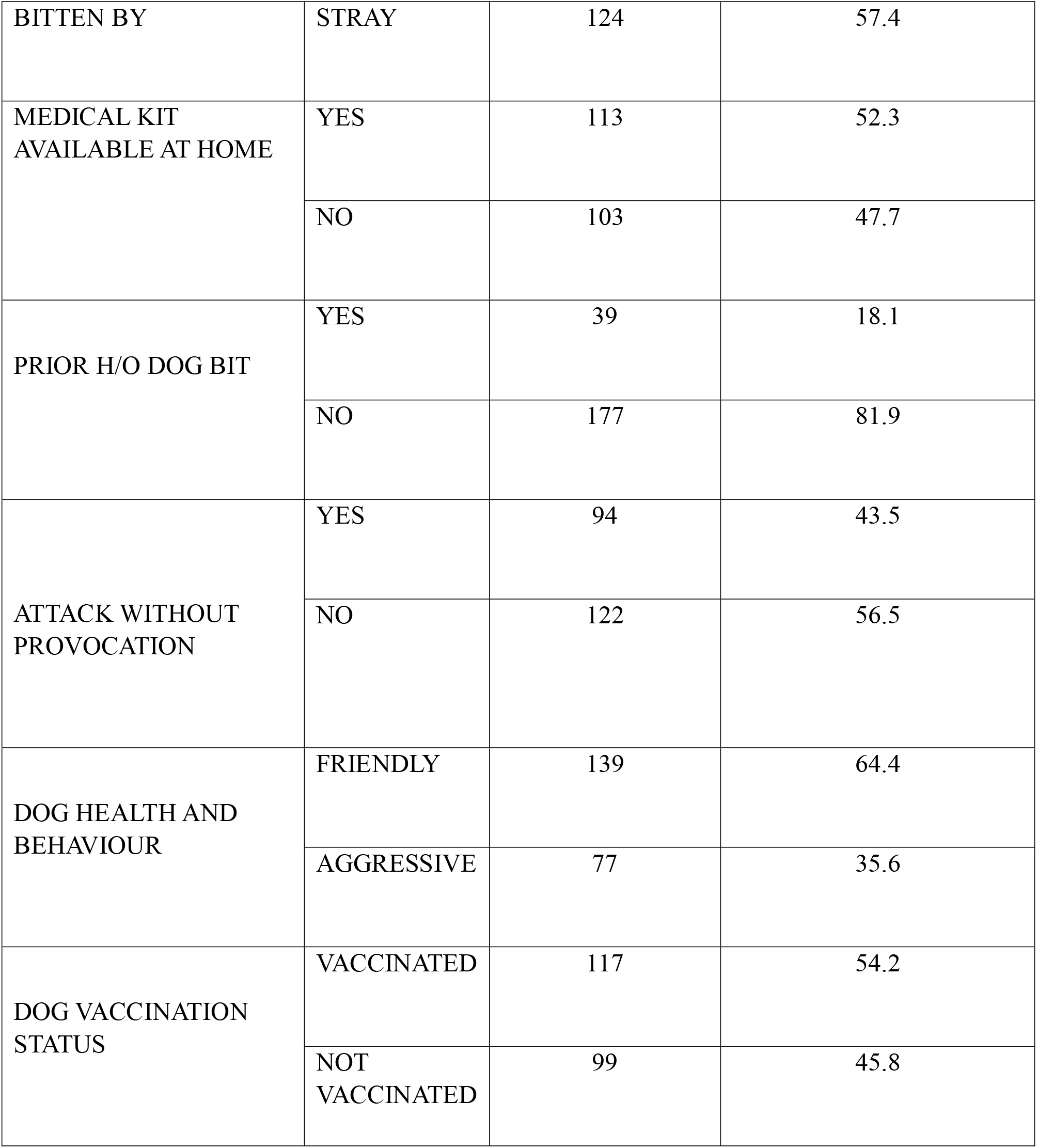
FREQUENCY AND PERCENTAGE DISTRIBUTION OF STUDY PARTICIPANTS BASED ON THE CIRCUMSTANCES OF DOG BITE EXPOSURE CHARACTERISTICS AND ASSOCIATED FACTORS (N=216)

**(TABLE -3)** The present study provides important insights into the management practices and treatment-seeking behaviour of animal bite cases in accordance with the World Health Organization (WHO) classification and the national guidelines for rabies prophylaxis. A total of 216 cases were analysed, revealing both strengths and gaps in current practices. In this study, 51.9% of patients were prescribed antibiotics, while 48.1% did not receive them. Antibiotic use in animal bite cases is generally indicated for high-risk wounds, such as deep injuries, bites on the hands, or wounds with a high risk of infection. The nearly equal distribution suggests a judicious but variable prescribing pattern, possibly influenced by clinical judgment and wound severity. However, without clear stratification by wound characteristics, there remains a possibility of both overuse and underuse, highlighting the need for standardized protocols.

**TABLE -3.** DISTRIBUTION OF MANAGEMENT OF DOG BITE INJURIES.

| VARIABLES |  | FREQUENCY<br>(N=216) | PERCENTAGE<br>% |
| --- | --- | --- | --- |
| WHO DOG<br>BITTENCriteria | CATEGORY I | 156 | 72.2 |
|  | CATEGORY II | 45 | 20.8 |
|  | CATEGORY III | 15 | 6.9 |
| ANTIBIOTIC PRESCRIBED | YES | 112 | 51.9 |
|  | NO | 104 | 48.1 |
| ANY FORM OF FIRST AID<br>GIVEN AT HOME | WASHED<br>WITH SOAP<br>AND WATER | 105 | 48.6 |
|  | CLEANED<br>WITH<br>ANTISEPTIC | 67 | 31.0 |
|  | APPLIED GEL<br>/ OINTMENT | 44 | 20.4 |
| TETANUS TOXOID<br>ADMINISTERED | YES | 185 | 85.6 |
|  | NO | 31 | 14.4 |
| ANTI-RABIS VACCINE<br>(COMPLETED FULL DOSE) | YES | 119 | 55.1 |
|  | NO | 97 | 44.9 |
| ADMITTED TO HOSPITAL<br>WARD | YES | 36 | 16.7 |
|  | NO | 180 | 83.3 |
| REFERRED TO SPECIALITY<br>CLINIC | YES | 56 | 25.9 |
|  | NO | 160 | 74.1 |

First aid practices at home showed that only 48.6% of patients washed their wounds with soap and water, which is the most effective immediate measure to reduce rabies transmission. A considerable proportion of patients relied on antiseptics (31.0%) or application of ointments (20.4%), which are not substitutes for proper wound washing. This indicates a significant gap in public awareness, as early and thorough washing of the wound is known to markedly decrease viral load and infection risk. These findings underscore the importance of community education programs focusing on appropriate first aid following animal bites.

Tetanus prophylaxis was administered in 85.6% of cases, reflecting good adherence to standard clinical guidelines. Given the risk of tetanus in open wounds, this high coverage is a positive indicator of appropriate preventive care. However, the remaining 14.4% who did not receive tetanus toxoid represent a missed opportunity for complete prophylaxis.

Regarding anti-rabies vaccination (ARV), only 55.1% of patients completed the full course, while 44.9% did not. This is a major concern, as incomplete vaccination leaves individuals at continued risk of developing rabies, a disease that is almost invariably fatal once clinical symptoms appear. Poor compliance may be attributed to factors such as lack of awareness, financial constraints, accessibility issues, or negligence, and it highlights the urgent need for better follow-up systems and patient counselling.

The majority of cases (83.3%) were managed on an outpatient basis, with only 16.7% requiring hospital admission, suggesting that most bites were mild or moderate in severity. Additionally, 25.9% of patients were referred to specialty clinics, indicating appropriate referral practices for complicated or high-risk cases. This reflects a functional tiered healthcare system, although further evaluation is needed to assess referral appropriateness.

According to WHO exposure classification, 72.2% of cases were Category I, followed by 20.8% Category II and 6.9% Category III. The predominance of Category I cases, which technically do not require post-exposure prophylaxis, suggests that a large number of patients sought medical care for low-risk exposures, possibly due to fear and lack of awareness. At the same time, nearly 28% of cases (Category II and III) required active prophylaxis, indicating a substantial burden of potentially rabies-exposing injuries.

An important observation is the discrepancy between exposure category and vaccination completion. Despite a high proportion of Category I cases, more than half of the total study population completed anti-rabies vaccination, suggesting either overutilization of vaccines or cautious clinical practice. While erring on the side of caution is understandable given the fatal nature of rabies, strict adherence to guidelines is essential to optimize resource utilization, especially in resource-limited settings.

Overall, the findings of this study highlight that while certain aspects of animal bite management such as tetanus prophylaxis and referral practices are satisfactory, there are critical gaps in first aid practices, vaccination compliance, and adherence to exposure-based treatment protocols. Strengthening public health education, improving accessibility to vaccines, and ensuring strict implementation of national guidelines are essential steps toward reducing the burden of rabies and improving patient outcomes.

**(TABLE -4)** The association between selected variables and the anatomical location of dog bite injuries was analysed. Among individuals aged less than 18 years, injuries were most commonly observed in the upper extremities (n = 25), followed by the lower extremities (n = 16), with fewer cases involving the head and neck (n = 2) and trunk (n = 1). Similarly, among those aged above 18 years, the lower extremities were most frequently affected (n = 106), followed by upper extremities (n = 57), trunk (n = 6), and head and neck (n = 3); however, this association was not statistically significant (p = 0.15).

**TABLE -4.**
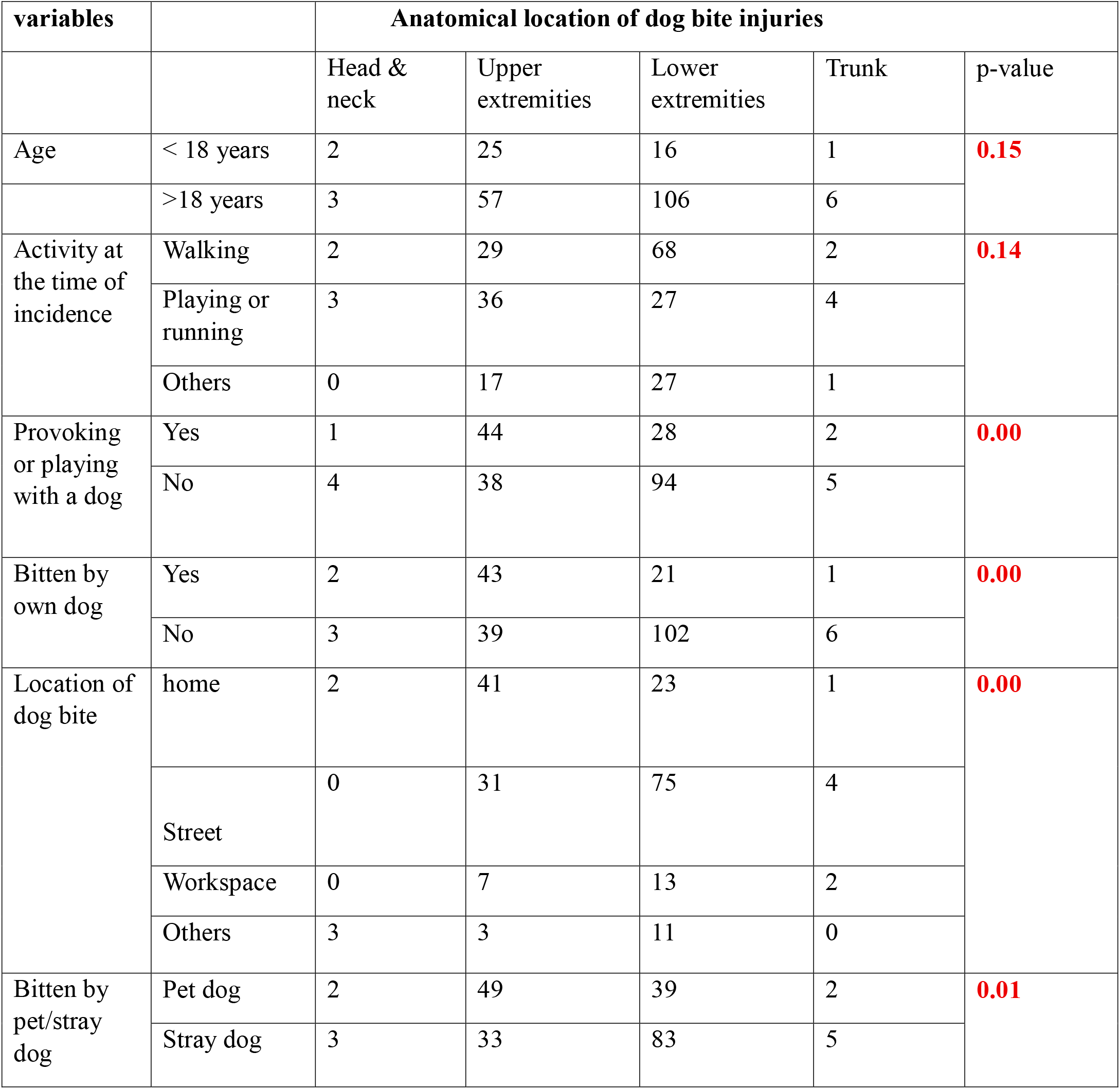
ASSOCIATION BETWEEN ANATOMICAL LOCATION OF DOG BITE INJURIES AND VARIOUS FACTORS.

With respect to activity at the time of incidence, individuals who were walking predominantly sustained injuries to the lower extremities (n = 68), whereas those playing or running had a relatively higher proportion of upper extremity injuries (n = 36) and lower extremity injuries (n = 27). Those engaged in other activities also showed a predominance of lower extremity involvement (n = 27). This association was not statistically significant (p = 0.14).

A statistically significant association was observed between provoking or playing with a dog and the anatomical site of injury (p = 0.00). Among those who reported provoking or playing with a dog, injuries were more common in the upper extremities (n = 44), whereas among those who did not, lower extremity injuries predominated (n = 94).

Similarly, being bitten by one’s own dog showed a significant association with injury location (p = 0.00). Individuals bitten by their own dog more frequently sustained upper extremity injuries (n = 43), while those bitten by other dogs had a higher proportion of lower extremity injuries (n = 102).

The location of the dog bite incident was also significantly associated with the anatomical site of injury (p = 0.00). Bites occurring at home were more commonly associated with upper extremity injuries (n = 41), whereas those occurring on the street predominantly affected the lower extremities (n = 75). Workplace and other locations also showed a higher frequency of lower extremity involvement.

Furthermore, the type of dog involved demonstrated a statistically significant association (p = 0.01). Bites from pet dogs were more frequently associated with upper extremity injuries (n = 49), whereas bites from stray dogs predominantly involved the lower extremities (n = 83).

**(TABLE -5)** The distribution of World Health Organization (WHO) bite exposure categories was analysed across different variables such as location of bite, type of dog, dog behaviour, number of biting attempts, and survival status of the dog.

**TABLE -5.** ASSOCIATED BETWEEN WHO BITE EXPOSURE CATEGORIES AND VARIOUS FACTORS.

**TABLE -4 ASSOCIATION BETWEEN ANATOMICAL LOCATION OF DOG BITE INJURIES AND VARIOUS FACTORS**
| variables |  | WHO Bite Exposure Categories |  |  | p-value |
| --- | --- | --- | --- | --- | --- |
|  |  | Categories 1 | Categories 2 | Categories 3 |  |
| Location of dog bite | Home | 58 | 8 | 1 | **0.05** |
|  | Street | 74 | 29 | 7 |  |
|  | Workspace | 13 | 4 | 5 |  |
|  | Others | 11 | 4 | 2 |  |
| Bitten by pet/stray dog | Pet dog | 78 | 12 | 2 | **0.01** |
|  | Stray dog | 78 | 13 | 13 |  |
| Dog health & behaviour | Friendly | 110 | 23 | 6 | **0.07** |
|  | Aggressive | 46 | 22 | 9 |  |
| Number of biting attempts | Single | 146 | 34 | 8 | **0.00** |
|  | Multiple | 10 | 11 | 7 |  |
| Dog survival status after bite | Alive | 126 | 30 | 8 | **0.11** |
|  | Dead | 1 | 0 | 1 |  |
|  | Unknown | 29 | 15 | 6 |  |

With regard to the location of dog bite, the majority of Category I bites occurred on the street (74 cases) and at home (58 cases), whereas Category II bites were also more common on the street (29 cases). Category III bites were highest in workplace settings (5 cases) and on the street (7 cases). This association was found to be statistically significant (p = 0.05), indicating that the location of the bite has a meaningful relationship with the severity of exposure.

In terms of type of dog, Category I bites were equally distributed between pet dogs (78 cases) and stray dogs (78 cases). However, Category III bites were notably higher among stray dogs (13 cases) compared to pet dogs (2 cases). This association was statistically significant (p = 0.01), suggesting that bites from stray dogs are more likely to result in severe exposure.

Regarding dog health and behaviour, most Category I bites were from friendly dogs (110 cases), while Category II and III bites were more frequently associated with aggressive dogs (22 and 9 cases, respectively). However, this association was not statistically significant (p = 0.07).

When analysing the number of biting attempts, the majority of Category I bites were due to a single bite (146 cases), whereas multiple bites were more commonly associated with higher severity categories, including Category II (11 cases) and Category III (7 cases). This relationship was found to be highly statistically significant (p = 0.00), indicating that multiple bite attempts significantly increase the risk of severe exposure.

Finally, considering the dog’s survival status after the bite, most cases involved dogs that were alive (126 Category I, 30 Category II, and 8 Category III cases). A small number of cases involved dogs that were dead or of unknown status. However, the association between dog survival status and exposure category was not statistically significant (p = 0.11).

## 4. DISCUSSION

The present study provides comprehensive insights into the epidemiology, clinical profile, and management of dog bite cases in accordance with the World Health Organization (WHO) classification and national rabies prophylaxis guidelines. The findings highlight important trends in demographic distribution, exposure patterns, and treatment practices, while also identifying critical gaps when compared with existing literature.

In the current study, the majority of victims were adults aged above 18 years (79.6%), with a male predominance (56.5%). This pattern is consistent with previous studies by which reported higher exposure among adult males due to increased outdoor activity and occupational exposure(Garg et al., 2013) However, some studies, such as (“ Antecedents and Consequences of Pediatric Dog-Bite Injuries and Their Developmental Trends,” n.d.)have demonstrated a higher incidence among children, particularly involving facial injuries, suggesting that age-related risk varies by setting and behaviour.

A higher proportion of cases originated from rural areas (60%), which aligns with findings from (Dutta et al., 2022), who reported increased vulnerability in rural populations due to limited awareness, poor access to healthcare, and higher stray dog populations. The early healthcare-seeking behaviour observed in this study, with 63% presenting within 24 hours, is encouraging and comparable to findings in other Indian studies, though delays beyond 24 hours still persist and remain a concern (Dutta et al., 2022).

In terms of exposure circumstances, most bites occurred while walking (46.8%) or during roadside activities, and a significant proportion were unprovoked (43.5%). Similar findings have been reported in global and regional studies, indicating that unprovoked bites constitute a major public health concern (*Animal Bites*, n.d.-b). Additionally, the predominance of street-based incidents (50.9%) and stray dog involvement (57.4%) is consistent with studies conducted in low- and middle-income countries, where free-roaming dog populations contribute significantly to rabies transmission (Acharya et al., 2026).

The anatomical distribution of injuries in this study showed that lower extremities were most commonly affected (56.5%), followed by upper extremities (38.0%). This is in agreement with studies by Duncan-Sutherland et al. (2022), which reported a similar predominance of lower limb injuries in adults. However, among children, upper extremity involvement was relatively higher, consistent with findings by(“ Antecedents and Consequences of Pediatric Dog-Bite Injuries and Their Developmental Trends,” n.d.), suggesting that behavioural differences influence injury patterns. The association analysis revealed that provoking or playing with dogs and being bitten by one’s own dog were significantly associated with upper extremity injuries. This aligns with previous literature indicating that closer human–animal interaction increases the likelihood of bites to accessible body parts such as hands and arms (Dixon et al., 2012). Similarly, bites from stray dogs and those occurring on the street were significantly associated with lower extremity injuries, reflecting typical defensive or territorial attacks.

Regarding severity, the majority of cases were classified as Category I (72.2%), followed by Category II (20.8%) and Category III (6.9%). This distribution is comparable to other studies in India, where minor exposures predominate, though a substantial proportion still requires post-exposure prophylaxis (*Bharadva: Epidemiology of Animal Bite Cases Attending*… *-Google Scholar*, n.d.). The significant association between multiple bite attempts and higher exposure categories observed in this study is consistent with clinical expectations and reinforces findings from other epidemiological studies.

Management practices in this study showed mixed adherence to guidelines. Antibiotics were prescribed in 51.9% of cases, similar to patterns reported in clinical reviews such as Stat Pearls, where antibiotic use is often guided by wound severity and infection risk (*Animal Bites - StatPearls - NCBI Bookshelf*, n.d.).

However, variability in prescription practices suggests the need for standardized treatment protocols. First aid practices were suboptimal, with only 48.6% of patients washing wounds with soap and water. This finding is consistent with studies by(Dutta et al., 2022)and (Beasley et al., 2022), which highlight poor awareness and reliance on traditional or inappropriate remedies in many communities. Given that immediate wound washing significantly reduces rabies transmission risk, this gap represents a major public health challenge.

Tetanus prophylaxis coverage was high (85.6%), reflecting good adherence to clinical guidelines. In contrast, completion of anti-rabies vaccination was observed in only 55.1% of cases, which is concerning. Similar issues of incomplete post-exposure prophylaxis have been widely reported in low-resource settings, often due to financial constraints, lack of follow-up, and limited awareness (Acharya et al., 2026). This underscores the need for improved health system strengthening and patient education.

The discrepancy between WHO exposure categories and vaccination practices suggests possible overutilization of vaccines in low-risk cases or cautious clinical decision-making, a phenomenon also noted in other studies. While caution is justified given the fatal nature of rabies, adherence to guidelines is essential to ensure optimal resource utilization.

Finally, the significant association between antibiotic prescription and immunization completion (p = 0.02) suggests that patients receiving more intensive clinical management are also more likely to adhere to preventive measures. This may reflect better counselling or increased perceived severity among these patients.

## 5. CONCLUSION

Dog bite injuries remain a significant yet preventable public health problem in Chennai. Despite a good proportion of patients presenting within 24 hours, gaps were identified in first-aid practices and rabies post-exposure prophylaxis uptake, with many victims not performing adequate wound washing or receiving complete immunization. Behavioural and environmental factors such as provocation, stray dog exposure, street incidents and multiple bite attempts were important predictors of injury location and severity.Overall, the findings highlight the urgent need for strengthened public health interventions including community awareness on immediate wound care and early medical consultation, improved access to rabies vaccination and post-exposure prophylaxis, responsible pet ownership and effective stray dog population control programmes.

## Data Availability

All data produced in the present study are available upon reasonable request to the authors

## ACKNOWLEDGMENTS

NONE

---

**FIGURE A.**
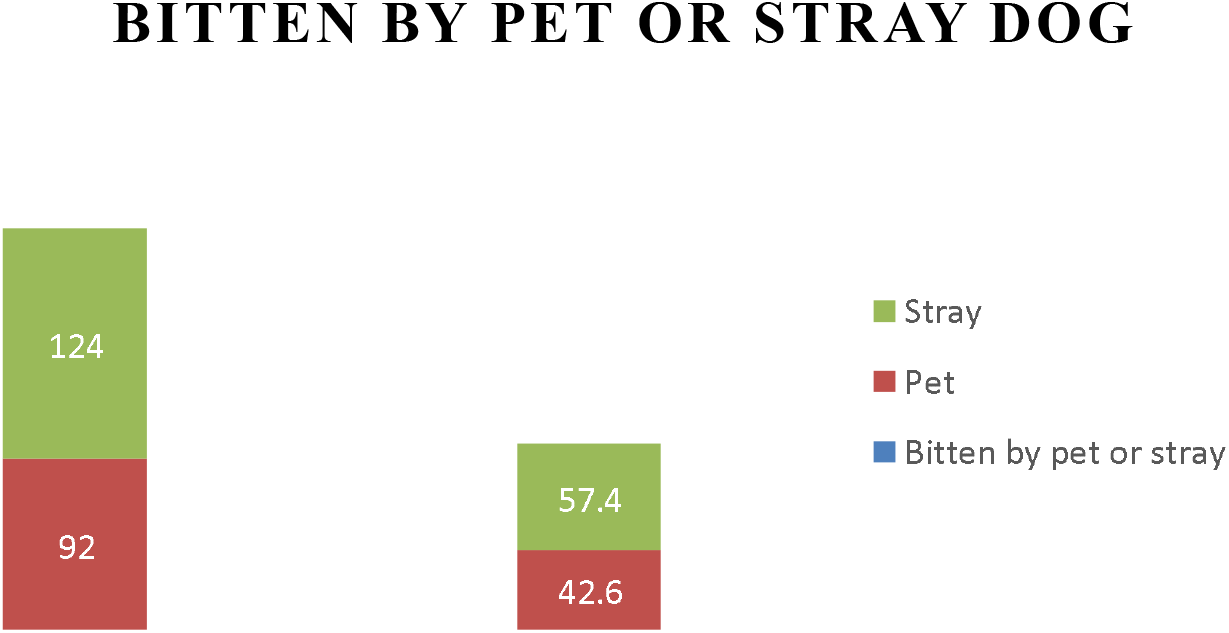
Most dog bites were caused by stray dogs, accounting for 57.4% (124 cases). Bites by pet dogs were less common, representing 42.6% (92 cases).

**FIGURE B.**
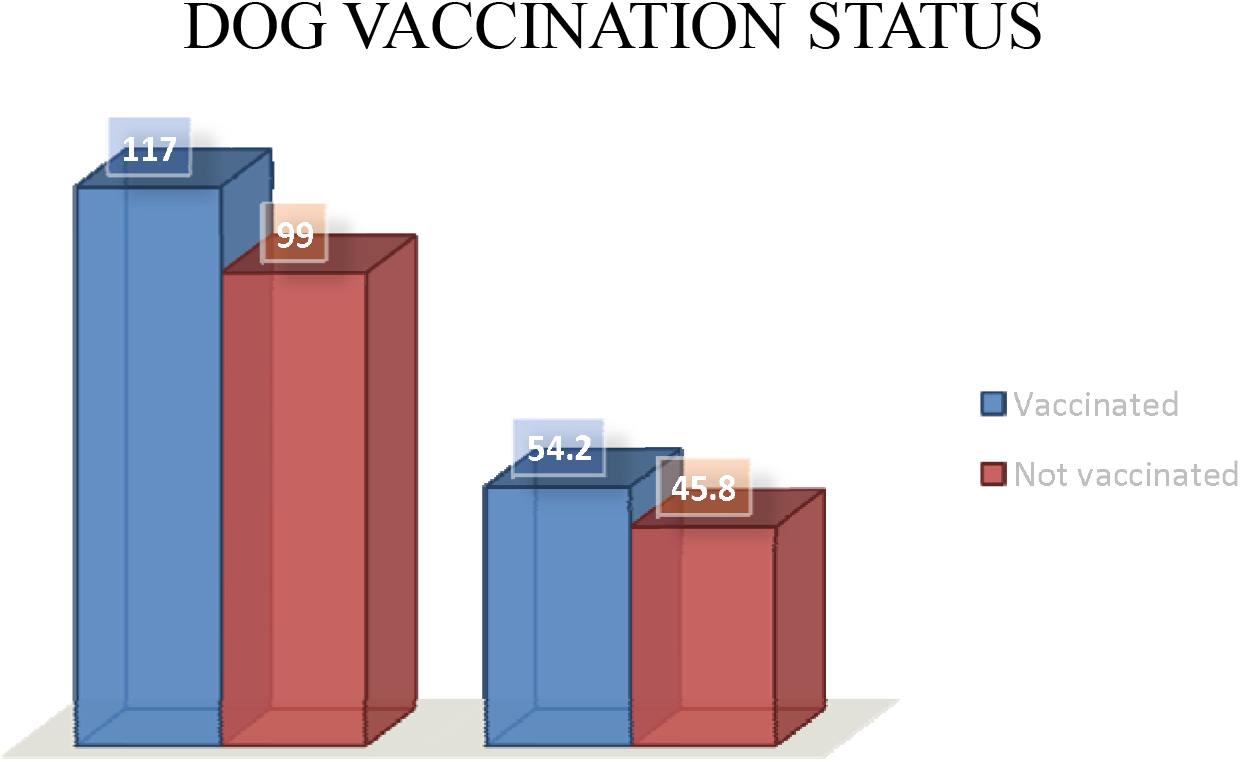
Out of 216 cases, 117 (54.2%) dogs were vaccinated and 99 (45.8%) dogs were not vaccinated. This indicates that slightly more than half of the cases of dog were vaccinated

**FIGURE C.**
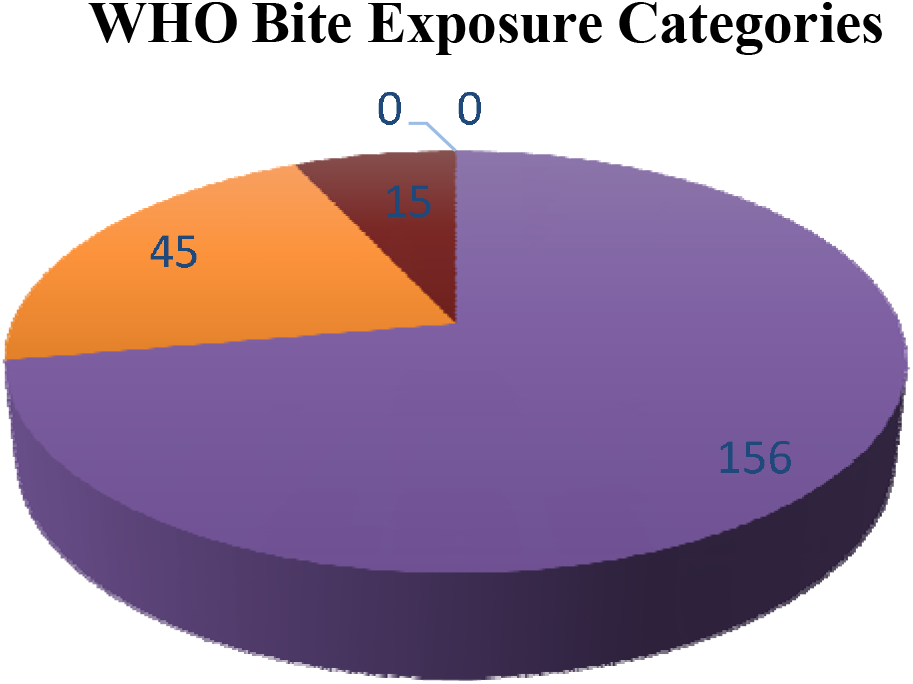
Out of 216 cases, most wounds were classified as Categories 1 (156 cases, 72.2%), followed by Categories 2 (45 cases, 20.8%), and Categories 3(15 cases, 6.9%). This indicates that the majority of injuries were minor, with only a small proportion being severe.

**FIGURE D.**
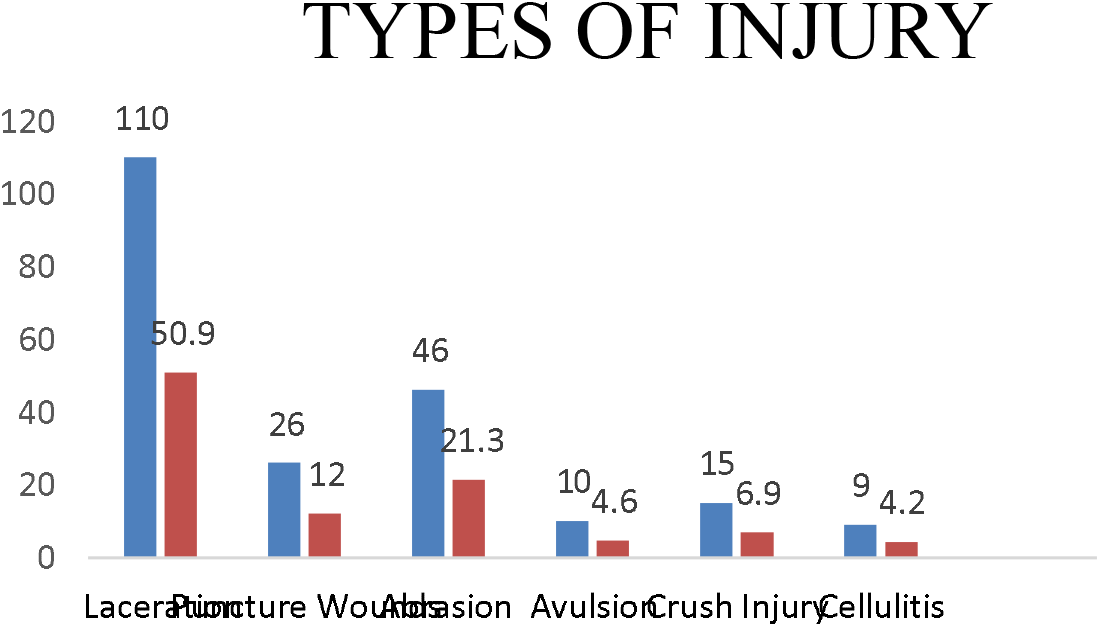
Out of 216 cases, lacerations were the most common type of injury(110 cases, 50.9%). This was followed by abrasions (46 cases, 21.3%) and puncture wounds (26 cases, 12.0%). Less common injuries included crush injuries (15 cases, 6.9%)

